# Methylenetetrahydrofolate reductase gene C677T polymorphism and risk of alcohol dependence

**DOI:** 10.1101/2020.06.15.20132332

**Authors:** Vandana Rai, Pradeep Kumar

## Abstract

Alcohol dependence is a complex neuropsychiatric disorder. Numerous studies investigated association between MTHFR gene C677T polymorphism and alcohol dependence (AD), but the results of this association remain conflicting. Accordingly, authors conducted a meta-analysis to further investigate such an association. PubMed, Elsevier Science Direct and Springer Link databases were searched for studies on the association between the MTHFR C677T polymorphism and AD. Pooled odds ratio (OR) with 95% confidence interval (CI) was calculated using the fixed- or random-effects model. Statistical analysis was performed with the software program MetaAnayst and MIX.

A total of 11 articles were identified through a search of electronic databases, up to February 28, 2020. The results of the present meta-analysis did not show any association between MTHFR C677T polymorphisms and AD risk (for T vs. C: OR = 1.04, 95% CI = 0.88-1.24; CT vs. CC: OR=1.02, 95%CI= 0.62-1.68; for TT + CT vs. CC: OR = 1.10, 95% CI = 0.94-1.29; for TT vs. CC: OR = 1.01, 95% CI = 0.66-1.51; for TT vs. CT + CC: OR = 0.97, 95% CI = 0.66-1.40). Results of subgroup analysis showed no significant association between MTHFR C677T polymorphism with AD in Asian as well as in Caucasian population. In conclusion, C677T polymorphism is not a risk factor for alcohol dependence.

## Introduction

Alcohol dependence or Alcoholism, also regarded as alcohol use disorder (AUD), is a complex and relapsing neuropsychiatric disorder (Koob 2003; Volkow et al., 2009). World Health Organization(WHO) reported that approximately 140 million individuals addicted to alcohol globally, resulting in to 2.5 million death each year (WHO,2011). AD is regarded as a “reward deficiency syndrome” that intemperately affects public health (Comings and Blum,2000; Parry et al., 2011). It has been found to be influenced by both genetic and environmental factors (Prescott and Kendler,1999; Kendler et al., 2007). Exact patho-physiological and molecular mechanism of AD is not known yet. However, molecular genetic studies support that multiple genes determine an individual"s predisposition to AD (Begleiter et al., 1999; Sokolov et al., 2003). Heritability of AD likely plays an important role in its development and is determined to be moderate to high (Goldman et al.,2005; Heath et al.,1997). It was reported frequently that alcohol consumption increased homocysteine (Hcy) concentration i.e hyperhomocysteinaemia (Bleich et al.,2005). However, inconsistent results of the combined effect of both positive and negative association have been reported between alcohol intake and Hcy (Ganji and Kafai 2003). Hyperhomocystenemia is already reported as risk factor for several diseases or disorders including neural tube defects, Alzheimer disease, schizophrenia, pregnancy complications, cardiovascular diseases, noninsulin dependent diabetes and end-stage renal disease as evidenced from several studies (Seshadri et al.,2002).

Homocysteine is a sulfur containing amino acid, several genetic and environmenta ris factor are reported for higher plasma concentration of homocysteine (Gudnason et al.,1998). Homocysteine (Hcy) is synthesized in methionine and folate cycle by demethylation of methionine. 5,10-methylenetetrahydrofolate reductase (MTHFR) enzyme of folate cycle plays an important role in homocysteine metabolism. MTHFR gene is present on chromosome 1p36.3. Numerous single nucleotide polymorphisms (SNP) are known in MTHFR gene like C677T and A1298C etc (Frosst et al. 1995; Goyette et al., 1995). The most clinically important and studies polymorphism is C677T (rs 1801133), in which cytosine (C) is substituted with thymine (T) at 677 nucleotide position and consequently alanine is replaced by Valine in MTHFR enzyme(Ala 222 Val) (Rozen et al., 1997; Chango et al., 2000). The variant MTHFR enzyme is thermolabile with reduced activity (‘∼70%) and it increased the plasma homocysteine concentrations (Frosst et al.,1995). Globally, frequency of mutant T allele varies greatly (Rady et al., 2002; Wilcken et al., 2003; Rai et al., 2010, 2012; Yadav et al., 2018). Yadav et al (2018) have conducted a comprehensive C667T polymorphism study and reported the highest frequency in European populations ranging from 24.1% to 64.3% and, lowest frequency from African population. Several studies revealed association of MTHFR gene C677T polymorphism with AD. However, findings showed inconsistent results (Lutz et al., 2006; Saffroy et al., 2008; Benyamina et al., 2009). To derive a more precise estimation of the relationship, authors performed a meta-analysis.

## Methods

Meta-analysis of observational studies in epidemiology (MOOSE) guidelines (Stroup et al., 2006) are followed in present meta-analysis.

### Retrieval strategy and selection criteria

Articles were retrieved through Pubmed, Google scholar, Springer Link, and Science Direct databases up to February 28, 2020, using following key words: ‘Methylenetetrahydrofoate reductase’ or ‘MTHFR’ or ‘C677T’ and ‘Alcohol dependence’ or ‘AD’ or ‘Addiction’.

### Inclusion and exclusion criteria

Inclusion criteria were following: (1) MTHFR C677T polymorphism and alcohol dependence association was investigated in the study, (2) MTHFR C677T genotype/ allele numbers in alcohol dependence cases and controls were given in the study and (3) sufficient information for calculating the odds ratio (OR) with 95% confidence interval (CI). Major reasons for studies exclusion were as follows: (1) no alcohol dependence cases analyzed, (2) the C677T polymorphism details information missing, and (3) duplicate article.

### Data extraction

Name of first author, country name, number of cases and controls, number of genotypes in cases and controls and journal name with full reference from each article were extracted.

### Statistical Analysis

All analysis were done according to the method of Rai et al (2014). ORs with 95 % confidence intervals (CIs) were calculated using fixed effect and random effect models (Mantel and Haenszel, 1959; Dersimonian and Laird, 1986). A five genetic models viz. alelle contrast, co-dominant, homozygote, dominant and recessive models were calculated. Heterogeneity was investigated and quantified by I^2^ statistic (Higgins and Thompson,2002). Chi-squared analysis was used to determine whether the genotype distribution of control group was in Hardy–Weinberg equilibrium or not. Subgroup analysis was conducted by ethnicity. Publication bias was investigated by Egger"s regression intercept test (Egger et al., 1997). P value <0.05 was considered statistically significant. All calculations were done by softwares MIX version 1.7 (Bax et al., 2006) and MetaAnalyst (Wallace et al., 2013) program.

## Results

### Eligible studies

Following the exclusion criteria, 10 individual case-control studies with a total of 1676 cases and 1594 controls were included into this meta-analysis (Bonsch et al., 2006; Lutz et al.,2006; Lutz et al., 2007; Saffroy et al., 2008; Benyamina et al., 2009; Fabris et al., 2009; Shin et al., 2010; Supic et al.,2010; Singh et al.,2014; Singh et al.,2015). One author (Supic et al.,2010), reported their data in to two categories, we included both set of data as different studies. Hence, total number of included studies in present meta-analysis is eleven (Tabel 1).

### Summary Statistics

Overall, eleven studies provided 1676/1594 cases/controls for MTHFR C677T polymorphism. The prevalence of C and T alleles in AD cases was 71.22% and 28.79% respectively. The percentage frequency of TT genotype among cases and controls was 9.43% and 12.98%, respectively whereas prevalence of CT heterozygote among AD cases was 38.72% and 39.21% in controls. The prevalence of CC homozygote among AD cases and controls was 51.85% and 47.80%, respectively. Genotypes were in Hardy-Weinberg equilibrium in all controls. In control group the percentage of C and T allele frequencies was 67.41% and 32.59% respectively (Figure 1).

**Figure 1.**
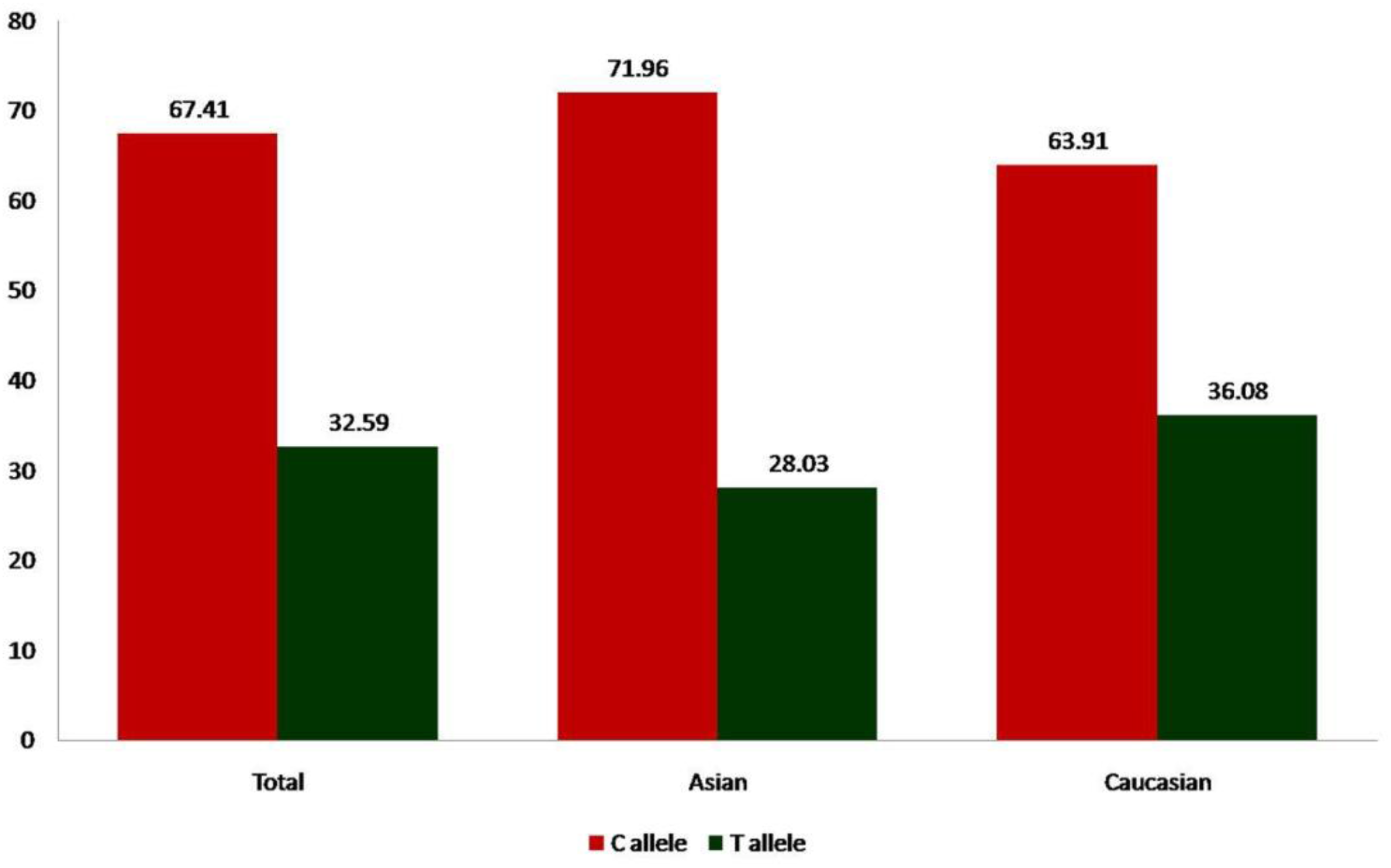
**Bar diagram showing percentage of C and T allele frequencies in control group of total 11 studies, 3 Asian studies and 8 Caucasian studies**.

### Meta-analysis

No significant association was observed between the MTHFR C677T polymorphism and the susceptibility to AD in all the genetic models using random effect model (for T vs. C (allele contrast): OR = 1.04, 95% CI = 0.88-1.24; CT vs. CC (co-dominant): OR=1.02, 95%CI= 0.62-1.68; for TT + CT vs. CC (dominant): OR = 1.10, 95% CI = 0.94-1.29; for TT vs. CC (homozygote): OR = 1.01, 95% CI = 0.66-1.51; for TT vs. CT + CC (recessive): OR = 0.97, 95% CI = 0.66-1.40)(Table 2; Figures 2).

**Table 1.** Details of included studies in the present meta-analysis.

| Study | Ethnicity | Control/Case | Case | HWE p-value |
| --- | --- | --- | --- | --- |
| Bonsch,2006 | Caucasian | 115 | 134 | 0.10 |
| Lutz,2006 | Caucasian | 102 | 221 | 0.98 |
| Lutz,2007 | Caucasian | 101 | 142 | 0.90 |
| Saffroy,2008 | Caucasian | 93 | 242 | 0.41 |
| Benyamina,2009 | Caucasian | 93 | 120 | 0.41 |
| Fabris,2009 | Caucasian | 236 | 63 | 0.55 |
| Shin,2010 | Asian | 232 | 68 | 0.07 |
| Supic,2010,Heavy Alcoholic | Caucasian | 57 | 32 | 0.84 |
| Supic,2010, Non heavy | Caucasian | 105 | 64 | 0.69 |

**Table 2.**

|  |  |  |  |  |
| --- | --- | --- | --- | --- |
| Alcoholic |  |  |  |  |
| Singh,2014 | Asian | 313 | 139 | 0.91 |
| Singh,2015 | Asian | 147 | 451 | 0.32 |

**Figure 2.**
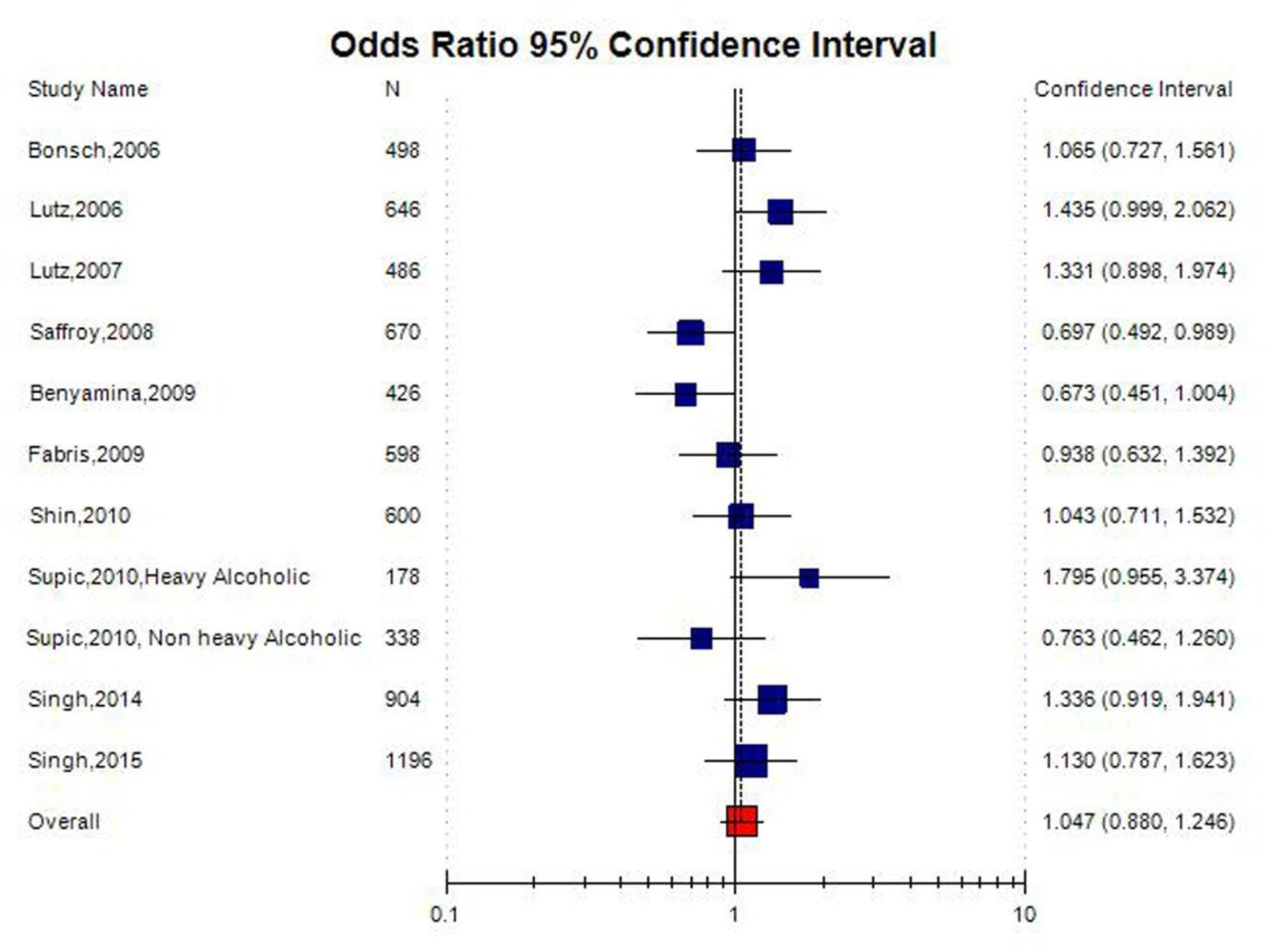
**Random effect Forest plot of allele contrast model (T vs. C) of total 11 studies of MTHFR gene C677T polymorphism**.

A true heterogeneity existed between studies for allele contrast (P_heterogeneity_ =0.02, Q= 20.64, I^2^= 51.56%, t^2^=0.04, z = 0.69), co-dominant genotype (P_heterogeneity_ <0.0001, Q= 86.64, I^2^=88.46%, t^2^=0.61, z = 4.29), homozygote genotype (P_heterogeneity_ =0.02, Q= 20.93, I^2^=52.24%, t^2^=0.24, z = 0.1), and recessive genotype (P_heterogeneity_ =0.02, Q= 21.00, I^2^= 52.40%, t^2^=0.20, z = 0.47) comparisons. The ‘I^2^’ value of more than 50% shows high level of true heterogeneity.

### Subgroup analysis

Out of 11 studies included in the present meta-analysis, 3 studies were carried out in Asian countries, and 8 studies were carried out on Caucasian (Table 1). The subgroup analysis by ethnicity did not reveal any significant association between MTHFR C677T polymorphism and AD in Asian population (T vs. C: OR= 1.16; 95% CI= 0.93-1.44; p= 0.17; I^2^= 3.1%; P_heterogeneity_= 0.65; TT vs. CC: OR= 1.16; 95% CI= 0.62-2.02; p= 0.69; I^2^= 3.1%; P_heterogeneity_= 0.89; and TT+CT vs. CC: OR= 1.26; 95% CI= 0.96-1.67; p= 0.09; I^2^= 3.1%; P_heterogeneity_=0.81); and Caucasian population (T vs. C: OR= 0.99; 95% CI= 0.86-1.14; p= 0.93; I^2^= 61.75%; P_heterogeneity_= 0.01; TT vs. CC: OR= 0.95; 95% CI= 0.70-1.29; p= 0.75; I^2^= 65.63%; P_heterogeneity_= 0.004; and TT+CT vs. CC: OR= 1.03; 95% CI= 0.85-1.25; p= 0.75; I^2^= 13.93%; Pheterogeneity=0.32)

### Publication bias

Symmetrical shape of Funnel plots’ revealed absence of pubication bias. P values of Egger’s test were more than 0.05, also provided statistical evidence for the funnel plots’ symmetry (p= 0.62for T vs. C; p= 0.48 for TT vs CC; p= 0.26 for CT vs. CC; p= 0.48 for TT+AC vs. CC; p= 0.28 for TT vs. CT+CC) (Table 3; Figure 4).

**Table 3:** Summary estimates for the odds ratio (OR) of MTHFR C677T in various allele/genotype contrasts, the significance level (p value) of heterogeneity test (Q test), and the I^2^ metric and publication bias p-value (Egger Test).

| **Genetic Models** | **Fixed effect**<br>**OR (95% CI), p** | **Random effect**<br>**OR (95% CI), p** | **Heterogeneity p-value (Q test)** | **$I^2$ (%)** | **Publication Bias (p of Egger's test)** |
| --- | --- | --- | --- | --- | --- |
| Allele Contrast (T vs. C) | 1.04 (0.92-1.17),0.48 | 1.04(0.88-1.24),0.60 | 0.02 | 51.56 | 0.62 |
| Dominant (TT+CT vs. CC) | 1.41(1.20-1.65),<0.0001 | 1.02(0.62-1.68),0.92 | <0.0001 | 88.46 | 0.06 |
| Homozygote (TT vs. CC) | 0.98(0.75-1.29),0.92 | 1.01(0.66-1.51),0.97 | 0.02 | 52.24 | 0.26 |
| Co-dominant (CT vs. CC) | 1.10(0.94-1.29),0.21 | 1.10(0.94-1.29),0.21 | 0.43 | 0 | 0.48 |
| Recessive (CC+CT vs. TT) | 0.94(0.73-1.20),0.63 | 0.97(0.66-1.40),0.86 | 0.02 | 52.4 | 0.28 |

**Figure 3.**
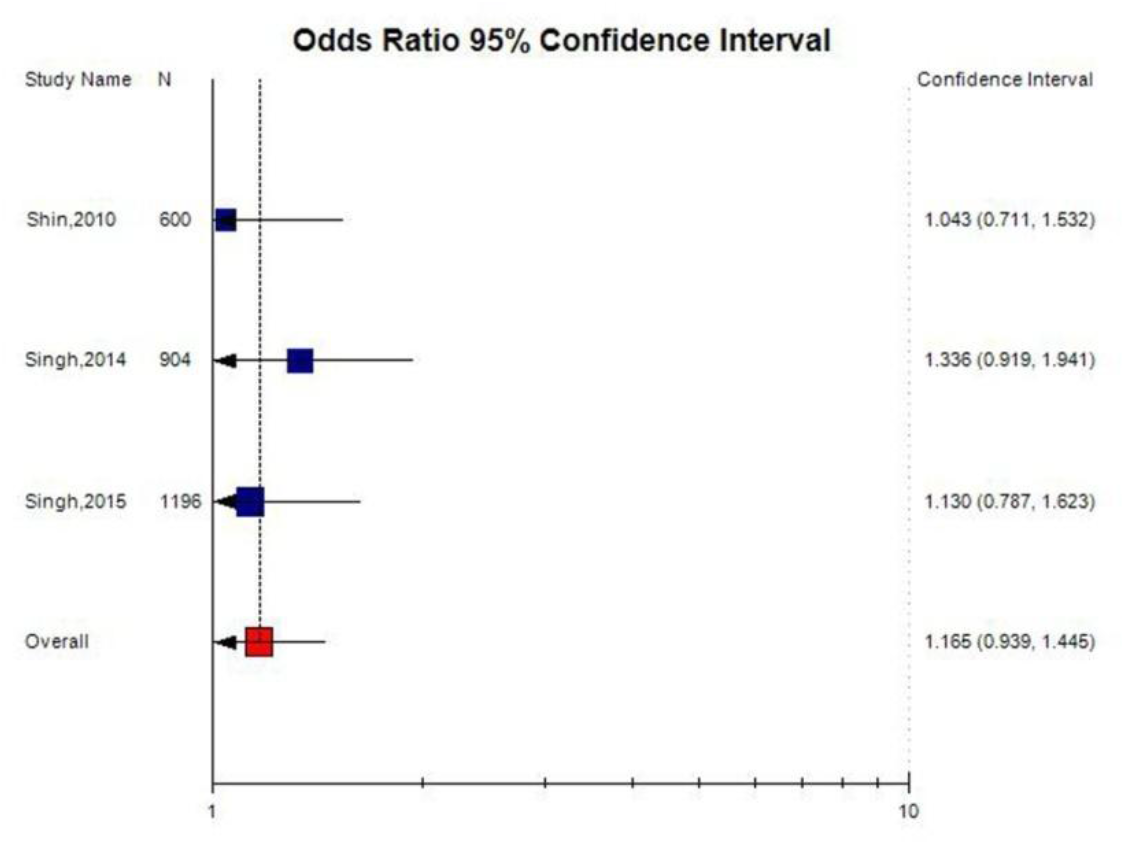
**Random effect Forest plot of allele contrast model (T vs. C) of total 3 Asian studies of MTHFR gene C677T polymorphism**.

**Figure 4.**
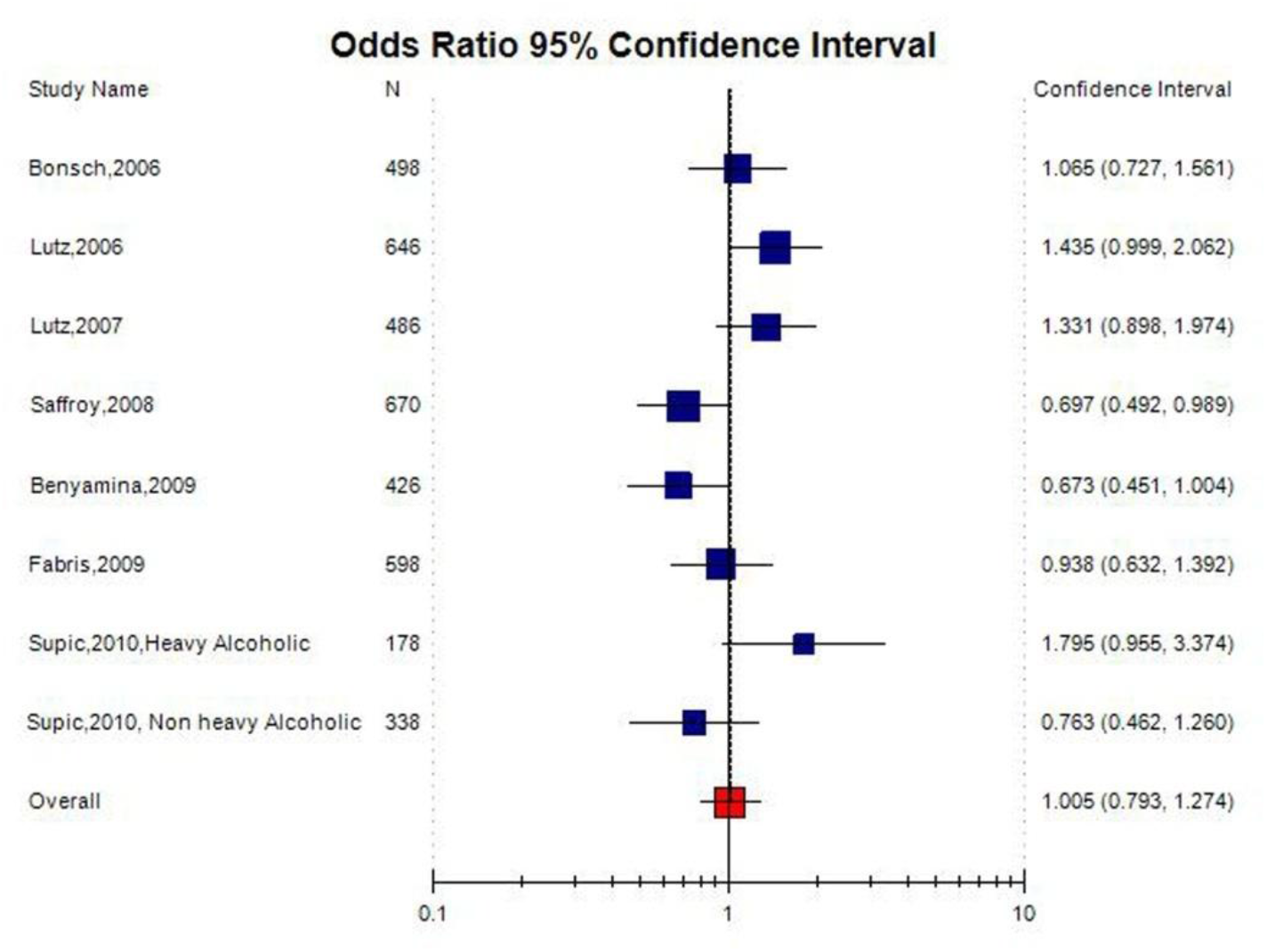
**Random effect Forest plot of allele contrast model (A vs. G) of total 8 Caucasian studies of MTHFR gene C677T polymorphism**.

**Figure 5.**
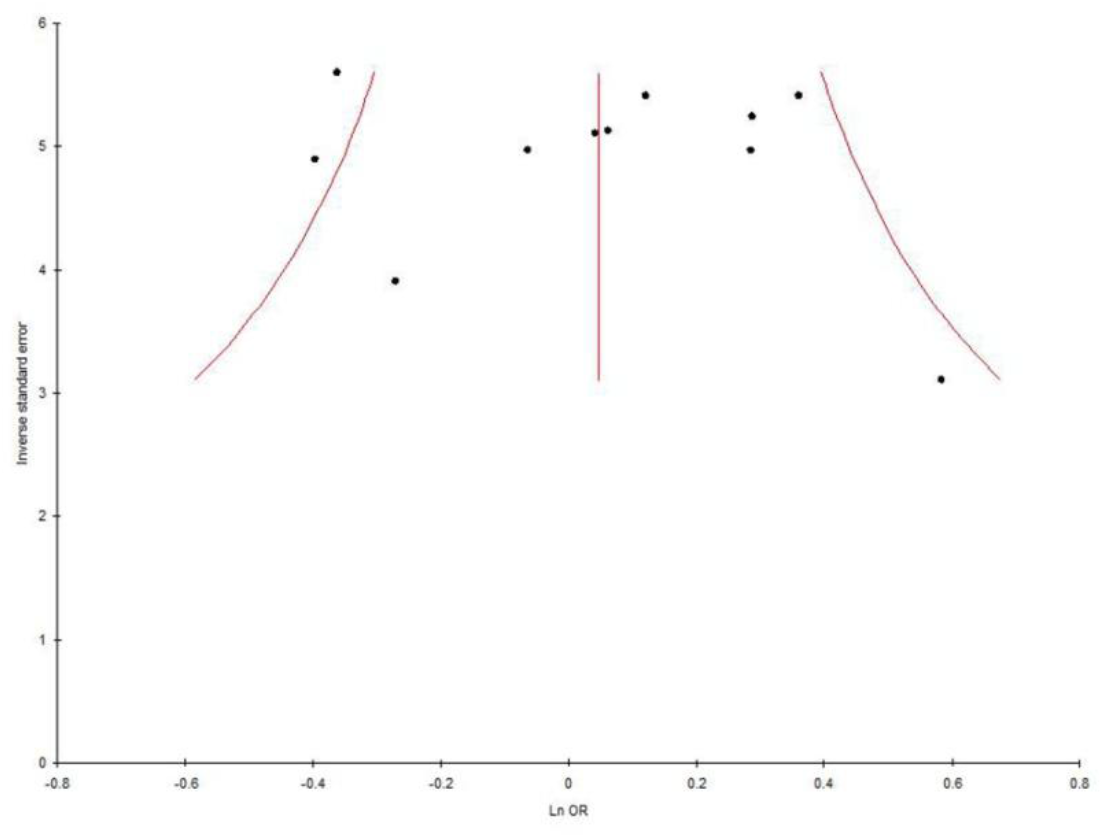
**Funnel plot-Precision by log odds ratio for allele contrast model (T vs. C) of total 11 studies of MTHFR gene C677T polymorphism**.

## Discussion

MTHFR enzyme is crucial for the methylation of homocysteine in to methionine and individulas with C677T polymorphism the plasma concentration of homocysteine is high. In vivo and in vitro studies has demonstrated that homocysteine has neurotoxic effects especially on dopamine neurons of reward pathway (Bleich et al. 2000). In addition, hyperhomocysteinemia is also reported in AD (Bleich et al 2000; Cravo and Camilo, 2000). According to deficit hypothesis of addiction, C677T polymorphism-dependent alteration of the reward system possibly leads to alcohol addiction.. Further, homovanillic acid (HVA) is a potential indicator of central dopaminergic neuronal activity (Amin et al., 1992) and experimentally, it was demonstrated that hyperhomocystein lowers the eve of HVA in rat stria region (Lee et al., 2005). On the basis of 11 studies providing data on MTHFR C677T genotype and AD risk in two ethnic populations, including over 3,205 subjects, our meta-analysis provides an evidence that TT and CT genotypes or T allele are not associated with AD risk. Hence the present meta-analysis indicated that C677T is a not a risk factor of AD.

Meta-analysis is a statistical tool to combine the information of independent case-control studies with similar target (Ioannidis et al., 2002). Several meta-analysis are published, which evaluated effects of gene polymorphisms in susceptibility of diseases/disorders-cleft lip and palate (Rai, 2017), Glucose-6-phosphate dehydrogenase deficiency (Kumar et al.,2016), down syndrome (Rai,2011; Rai et al., 2017; Rai and Kumar, 2018), male infertility (Rai and Kumar,2017), schizophrenia (Yadav et al., 2015; Rai et al., 2017), obsessive compulsive disorder (Kumar and Rai,2019), depression (Rai,2014,2017), epilepsy (Rai and Kumar,2018), Alzheimers disease (Rai, 2016), esophageal cancer (Kumar and Rai, 2018), prostate cancer (Yadav et al.,2016), breast cancer (Rai et al.,2017), digestive tract cancer (Yadav et al.,2018), ovary cancer (Rai, 2016), endometrial cancer (Kumar et al., 2018), uterine leiomyioma (Kumar and Rai, 2018), and MTRR gene frequency (Yadav et al.,2019).

Several limitations that should be acknowledged like (i)calculated crude Odds ratio, (ii) included the less number of available studies(10 studies) and the limited sample size of each included study, (iii) observed higher between study heterogeneity, (iv) considered only one gene polymorphism and (v) not considered other confounding factors like diet, gender etc. In addition to limitations, current meta-analysis has several strength also such as - higher study power and larger sample size in comparison to individual case control studies, and absence of publication bias etc.

In conclusion, pooled analysis of data from 11separate articles indicates that the MTHFR 677TT genotype is not a risk factor for AD. The results of present meta-analysis should be interpreted with certain cautions due to presence of higher heterogeneity and small number of included studies. Future large-scale, population-based association studies from different regions of the world are required to investigate potential gene–gene and gene–environment interactions involving the MTHFR C677T polymorphism in determining AD risk.

## Data Availability

Yes data are availabe

## References

Amin F, Davidson M. Davis KL. 1992. Homovanillic acid measurement in clinical research: A review of methodology. Schizophrenia Bulletin, 18(1):123–148.

Bax L, Yu LM, Ikeda N, Tsuruta H, & Moons KG. (2006). Development and validation of MIX: comprehensive free software for meta-analysis of causal research data. BMC Med Res Methodol. 6:50–58.

Begleiter H, Reich T, Nurnberger J, Li TK, Conneally PM, H. Edenberg H, R. Crowe R, Kuperman S, Schuckit M, Bloom F, Hesselbrock V, Porjesz B, Cloninger CR, Rice J, Goate A. Description of the Genetic Analysis Workshop 11 Collaborative Study on the Genetics of Alcoholism, Genetic epidemiology 17 Suppl 1 (1999) S25–30.

Benyamina A, Saffroy R, Blecha L, Pham P, Karila L, Debuire B, Lemoine A, Reynaud M. Association between MTHFR 677C-T polymorphism and alcohol dependence according to Lesch and Babor typology, Addiction biology 14 (2009) 503–505.

Bleich S, Carl M, Bayerlein K. et al., “Evidence of increased homocysteine levels in alcoholism: the Franconian alcoholism research studies (FARS),” Alcoholism: Clinical and Experimental Research, vol. 29, no. 3, pp. 334–336, 2005.

Bonsch D, Bayerlein K, Reulbach U, Fiszer R, Hillemacher T, Sperling W, Kornhuber J, Bleich S. Different allele-distribution of mthfr 677 C -> T and mthfr −393 C -> a in patients classified according to subtypes of Lesch’s typology, Alcohol and alcoholism 41 (2006) 364–367.

Chango A, Boisson F, Barbe F, Quilliot D, Droesch S, Pfister M, et al. The effect of 677C?T and 1298A?C mutations on plasma homocysteine and 5,10-methylenetetrahydrofolate reductase activity in healthy subjects. Br J Nutr. 2000;83:593–6.

Comings DE, Blum K. Reward deficiency syndrome: genetic aspects of behavioral disorders, Progress in brain research 126 (2000) 325–341.

Cravo ML, Camilo ME. Hyperhomocysteinemia in chronic alcoholism: relations to folic acid and vitamins B(6) and B(12) status. Nutrition. 2000;16(4):296–302.

DerSimonian R, Laird N. (1986).Meta-analysis in clinical trials. Control Clin Trials. 7:177–88.

Egger M, Smith GD, Schneider M, & Minder C. (1997). Bias in meta-analysis detected by a simple, graphical test. BMJ 315:629–634.

Fabris C, Pierluigi Toniutto, Edmondo Falleti, Elisabetta Fontanini, Annarosa Cussigh, Davide Bitetto, Ezio Fornasiere, Elisa Fumolo, Claudio Avellini, Rosalba Minisini, and Mario Pirisi. 2009. MTHFR C677T Polymorphism and Risk of HCC in Patients With Liver Cirrhosis: Role of Male Gender and Alcohol Consumption. Alcoholism: Clinical and Experimental Research 33, 102–107.

Frosst P, Blom HJ, Milos R, Goyette P, Sheppard CA, Matthews RG, et al. A candidate genetic risk factor for vascular disease: a common mutation in methylenetetrahydrofolate reductase. Nat Genet. 1995;10:111–3.

Ganji V, Kafai MR.2003.Demographic, health, lifestyle, and blood vitamin determinants of serum total homocysteine concentrations in the third National Health and Nutrition Examination Survey, 1988–1994. The American Journal of Clinical Nutrition, 77, 826–833.

Goldman D, Oroszi G, Ducci F. The genetics of addictions: uncovering the genes, Nature reviews. Genetics 6 (2005) 521–532.

Goyette P, Pai A, Milos R, Frosst P, Tran P, Chen Z, et al. Gene structure of human and mouse methylenetetrahydrofolate reductase (MTHFR). Mamm Genome. 1998;9:652–6.

Gudnason V, Stansbie D, Scott J, Bowron A, Nicaud V, Humphries S. C677T (thermolabile alanine/valine) polymorphism in methylenetetrahydrofolate reductase (MTHFR): its frequency and impact on plasma homocysteine concentration in different European populations. Atherosclerosis 136 (1998) 347–354.

Heath AC, Bucholz KK, Madden PA,. Dinwiddie SH, Slutske WS,. Bierut LJ, Statham DJ, Dunne MP, Whitfield JB, Martin NG. Genetic and environmental contributions to alcohol dependence risk in a national twin sample: consistency of findings in women and men, Psychological medicine 27 (1997) 1381–1396.

Higgins JP, Thompson SE. (2002). Quantifying heterogeneity in a meta-analysis. Stat Med. 21:1539–58.

Ioannidis JP, Rosenberg PS, Goedert JJ, & O’Brien TR. (2002). International meta-analysis of HIV host genetics. Commentary: meta-analysis of individual participants? data in genetic epidemiolog. Am J Epidemiol. 156: 204–210.

Kendler KS, Myers J, Prescott CA. Specificity of genetic and environmental risk factors for symptoms of cannabis, cocaine, alcohol, caffeine, and nicotine dependence, Archives of general psychiatry 64 (2007) 1313–1320.

Koob GF. Alcoholism: allostasis and beyond. Alcohol Clin Exp Res. 2003 ;27:232–243.

Kumar P, Yadav U, Rai V. (2016). Prevalence of glucose 6-pohsphate dehydrogenase deficiency in India: An updated meta-analysis. Egypt J Med Hum Genet. 2016; 17: 295–302.

Kumar P, Rai V. MTHFR C677T polymorphism and risk of esophageal cancer: An updated meta-analysis. Egyptian Journal of Medical Human Genetics. 2018; 19: 273–284.

Kumar P, Singh G, Rai V. (2020). Evaluation of COMT Gene rs4680 polymorphism as a risk factor for endometrial cancer. Ind J Clin Biochem. 35(1):63–71.

Kumar P, Rai V. (2018).Catechol-O-Methyltransferase Val158Met polymorphism and susceptibility to Uterine Leiomyoma. Jacobs Journal of Gynecology and Obstetrics,5(1): 043.

Kumar P, Rai V. (2020). Catechol-O-methyltransferase gene Val158Met polymorphism and obsessive compulsive disorder susceptibility: a meta-analysis. Metab. Brain Dis. 35:242-251.ISSN 0885-7490.

Lee EY, Hongtao Chen,1 Karam F.A. Soliman,1 Clivel G. Charlton. 2005. Effects of Homocysteine on the Dopaminergic System and Behavior in Rodents. NeuroToxicology 26 (2005) 361–371.

Lutz UC, Batra A, Kolb W, Machicao F, Maurer S, Kohnke MD. Methylenetetrahydrofolate reductase C677T-polymorphism and its association with alcohol withdrawal seizure, Alcoholism, clinical and experimental research 30 (2006) 1966–1971.

Lutz UC, Batra A, Wiatr G, Machicao F, Kolb W, Maurer S, Buchkremer G, Kohnke MD. Significant impact of MTHFR C677T polymorphism on plasma homovanillic acid (HVA) levels among alcohol-dependent patients, Addiction biology 12 (2007) 100–105.

Mantel N, and Haenszel W. 1959. Statistical aspects of the analysis of data from retrospective studies of disease. J Natl Cancer Inst. 22(4): 719–48.

Parry CD, Patra J, Rehm J. Alcohol consumption and non-communicable diseases: epidemiology and policy implications, Addiction 106 (2011) 1718–1724.

Prescott CA, Kendler KS. Genetic and environmental contributions to alcohol abuse and dependence in a population-based sample of male twins, The American journal of psychiatry 156 (1999) 34–40.

Rady PL, Szucs S, Grady J, Hudnall SD, Kellner LH, Nitowsky H, et al. Genetic polymorphisms of methylenetetrahydrofolate reductase (MTHFR) and methionine synthase reductase (MTRR) in ethnic populations in Texas; a report of a novel MTHFR polymorphic site, G1793A. Am J Med Genet. 2002;107:162–8.

Rai V, Yadav U, Kumar P, Yadav SK (2010) Methylenetetrahydrofolate Reductase Polymorphism (C677T) in muslim population of Eastern Uttar Pradesh, India. Indian J Med Sci 64(5): 219–23.

Rai V, Yadav U, Kumar P (2012) Prevalence of methylenetetrahydrofolate reductase C677T polymorphism in eastern Uttar Pradesh. Indian J Hum Genet 18(1): 43–6.

Rai V. (2011). Polymorphism in folate metabolic pathway gene as maternal risk factor for Down syndrome. Int J Biol Med Res., 2(4): 1055–1060.

Rai V. (2014). Genetic polymorphisms of methylenetetrahydrofolate reductase (MTHFR) gene and susceptibility to depression in Asian population: a systematic meta-analysis. Cell Mol Biol. 60 (3): 29–36.

Rai V, Yadav U, Kumar P, Yadav SK, Mishra OP. 2014.Maternal Methylenetetrahydrofolate Reductase C677T Polymorphism and Down Syndrome Risk: A Meta-Analysis from 34 Studies. PLOS ONE 9 e108552.

Rai V. (2016). Folate pathway gene methylenetetrahydrofolate reductase C677T polymorphism and Alzheimer disease risk in Asian population. Indian J Clin Biochem. 31(3): 245–52.

Rai V. 2016. Methylenetetrahydrofolate Reductase Gene C677T Polymorphism and Its Association with Ovary Cancer. J Health Med Informat 7:3

Rai V. Strong association of C677T polymorphism of methylenetetrahydrofolate reductase gene with nosyndromic cleft lip/palate (nsCL/P). Ind J Clin Biochem. 2017; 1–11.

Rai V, Yadav U, Kumar P, Yadav SK, Gupta S. (2017). Methylenetetrahydrofolate Reductase A1298C Genetic Variant and Risk of Schizophrenia: an updated meta-analysis. Indian J Med Res, 145(4):437–447.

Rai V, Yadva U, Kumar P. (2017). Null association of maternal MTHFR A1298C polymorphism with Down syndrome pregnancy: An updated meta-analysis. The Egyptian J Med Hum Genet. 18(1): 9–18.

Rai V. (2017). Association of C677T polymorphism (rs1801133) in MTHFR gene with depression. Cell Mol Biol, 2017;63(6):60–67.

Rai V, Yadav U, Kumar P. (2017). Impact of Catechol-O-Methyltransferase Val 158Met (rs4680) Polymorphism on breast cancer Susceptibility in Asian population. Asian Pac J Cancer Prev. 18 (5): 1243–1250.

Rai V, Kumar P. (2017). Methylenetetrahydrofolate Reductase C677T Polymorphism and Risk for Male Infertility in Asian Population. Ind J Clin. Biochem. 32(3), 253–260.

Rai V, Kumar P. (2018). Fetal MTHFR C677T polymorphism confers no susceptibility to Down Syndrome: evidence from meta-analysis. Egyptian J Med Hum Genet.19: 53–58.

Rai V, Kumar P. (2018). Methylenetetrahydrofolate reductase C677T polymorphism and susceptibility to epilepsy. Neurological Sciences https://doi.org/10.1007/s10072-018-3583-z.

Rozen R. Genetic predisposition to hyperhomocysteinemia: deficiency of methylenetetrahydrofolate reductase (MTHFR), Thrombosis and haemostasis 78 (1997) 523–526.

Saffroy R, Benyamina A, Pham P, Marill C, Karila L, Reffas M, Debuire B, Reynaud M, Lemoine A. Protective effect against alcohol dependence of the thermolabile variant of MTHFR, Drug and alcohol dependence 96 (2008) 30–36.

Seshadri S, Beiser A, Selhub J. et al., Plasma homocysteine as a risk factor for dementia and Alzheimer’s disease The New England Journal of Medicine, 346, no. 7, 476–483, 2002.

Shin S, Stewart R, Ferri CP, Kim JM, Shin I S, Kim SW, Yang S J, Yoon J S. 2010, An investigation of associations between alcohol use disorder and polymorphisms on ALDH2, BDNF, 5-HTTLPR, and MTHFR genes in older Korean men, International journal of geriatric psychiatry, 25 441–448.

Singh H K, Salam K, Saraswathy K N. 2014. A Study on MTHFR C677T Gene Polymorphism and Alcohol Dependence among Meiteis of Manipur, India, Journal of Biomarkers, Article ID 310241, 5 pages,http://dx.doi.org/10.1155/2014/310241.

Singh HS, Devi S, Saraswathy K. 2015. Methylenetetrahydrofolate reductase (MTHFR) C677T gene polymorphism and alcohol consumption in hyperhomocysteinaemia: a population-based study from northeast India. Journal of Genetics, 94 (1), 121–124.

Sokolov BP, Jiang L, Trivedi NS, Aston C, Transcription profiling reveals mitochondrial, ubiquitin and signaling systems abnormalities in postmortem brains from subjects with a history of alcohol abuse or dependence, Journal of neuroscience research 72 (2003) 756–767.

Stroup DF, Berlin JA, Morton SC, Olkin I, Williamson GD, & Rennie D, et al. (2000) Meta-analysis of observational studies in epidemiology: a proposal for reporting. Meta-analysis Of Observational Studies in Epidemiology (MOOSE) group. JAMA. 2000; 283(15): 2008–2012.

Supic G, Jovic N, Kozomara R, Zeljic K, Magic Z. 2011. Interaction Between the MTHFR C677T Polymorphism and Alcohol--Impact on Oral Cancer Risk and Multiple DNA Methylation of Tumor-Related Genes. J Dent Res. 90(1): 65–70.

Volkow ND, Fowler JS, Wang GJ, Baler R, Telang F. Imaging dopamine’s role in drug abuse and addiction. Neuropharmacology. 2009; 56 :3–8

Wallace BC, Dahabreh IJ, Trikalinos TA, Lau J, Trow P, Schmid CH. (2013). Closing the gap between methodologists and end-users: R as a computational back-end. J Stat Softw. 49: 1–15.

Wilcken B, Bamforth F, Li Z, Zhu H, Ritvanen A, Renlund M, et al. Geographical and ethnic variation of the 677C[T allele of 5,10 methylenetetrahydrofolate reductase (MTHFR): findings from over 7000 newborns from 16 areas world-wide. J Med Genet. 2003;40:619–25.

WHO, Global Status Report on Alcohol and Health, WHO, Geneva, Switzerland, 2011.

Yadav U, Kumar P, Gupta S, Rai V. (2016).Role of MTHFR C677T gene polymorphism in the susceptibility of schizophrenia: An updated meta-analysis. Asian J Psychiatry. 20: 41–51.

Yadav U, Kumar P, Rai V. (2016). Role of MTHFR A1298C gene polymorphism in the etiology of prostate cancer:a systematic review and updated meta-analysis. Egyptian Journal Medical Human Genetics. 17(2): 141–148.

Yadav U, Kumar P, Gupta S, Rai V. (2018). Distribution of MTHFR C677T Gene Polymorphism in Healthy North Indian Population and an Updated Meta-analysis. Ind J Clin Biochem. 32(4), 399–410. ISSN: 0974-0422.

Yadav U, Kumar P, Rai V. (2018).NQO1 gene C609T polymorphism (dbSNP: rs1800566) and digestive tract cancer risk: A Meta analysis. Nutrition and Cancer. 10.1080/01635581.20.

Yadav U, Kumar P, & Rai V. (2019). Distribution of Methionine Synthase Reductase (MTRR) Gene A66G Polymorphism in Indian Population. Ind J Clin Biochem. ISSN: 0974-0422. https://doi.org/10.1007/s12291-019-00862-9.

